# Estimating Demand for Potential Disease modifying Therapies for Alzheimer’s Disease in the UK

**DOI:** 10.1101/2023.11.17.23298682

**Authors:** Axel AS Laurell, Ashwin V Venkataraman, Tatjana Schmidt, Marcella Montagnese, Christoph Mueller, Robert Stewart, Jonathan Lewis, Clare Mundell, Jeremy D Isaacs, Mani S Krishnan, Robert Barber, Timothy Rittman, Benjamin R Underwood

## Abstract

**Background:** Phase three trials of the monoclonal antibodies lecanemab and donanemab, which target brain amyloid, have reported statistically significant differences in clinical endpoints in early Alzheimer’s disease. These drugs are already in use in some countries and are going through the regulatory approval process for use in the UK. Concerns have been raised about the ability of healthcare systems, including those in the UK, to deliver these treatments considering the resources required for their administration and monitoring.

**Aim:** To estimate the scale of real-world demand for monoclonal antibodies for AD in the United Kingdom.

**Method:** We used anonymised patient record databases from two National Health Service trusts for the year 2019 to collect clinical, demographic, cognitive and neuroimaging data for these cohorts. Eligibility for treatment was assessed using the inclusion criteria from the clinical trials of donanemab and lecanemab with consideration given to diagnosis, cognitive performance, cerebrovascular disease, and willingness to receive treatment.

**Results:** We examined the records of 82,386 people referred to services covering some 2.2 million people. After applying the trial criteria, we estimate that a maximum of 906 people per year would start treatment with monoclonal antibodies in the two services, equating to 30,200 people if extrapolated nationally.

**Conclusions:** Monoclonal antibody treatments for AD are likely to present a significant challenge for healthcare services to deliver in terms of the neuroimaging and treatment delivery. The data provided here allows health services to understand the potential demand and plan accordingly.

## Introduction

More than a century since Alzheimer’s disease (AD) was first described, the first phase 3 clinical trials demonstrating a statistically significant clinical response combined with changes in disease biomarkers have been published (1,2). These trials used donanemab and lecanemab, which are monoclonal antibodies targeting beta amyloid, rejuvenating interest in the potential of this approach. After many negative trials of monoclonal antibodies, the contentious re-appraisal of data from two large scale trials of aducanumab led researchers to believe that optimal design and dosing of anti-amyloid therapies might be associated with a modest clinical benefit (3). This led to the Food and Drug Administration’s (FDA) “accelerated approval” for aducanumab in the United States (USA) based largely on clinical extrapolation of reductions in brain amyloid, although the data for clinical outcomes were questionable. This was followed by the publication of a similar phase 2 trial of donanemab (TRAILBLAZER-ALZ (4)) and subsequently the phase 3 trials of lecanemab (CLARITY-AD (1)) and donanemab (TRAILBLAZER-ALZ 2 (2)) which more definitively reported slowing of decline in cognitive, functional, and other outcomes. The FDA approved lecanemab (marketed in the USA as Leqembi) under the accelerated approval process in January 2023 and gave it full FDA approval in July 2023 (5). Eisai and Biogen have subsequently submitted both a supplemental biologics licence application to the FDA and a marketing authorisation application to the European Medicines Agency (EMA).

Whether the magnitude of the demonstrated benefit justifies the risks of treatment and economic cost remains contentious (6,7). One concern has been the ability of healthcare systems, including those in developed nations like the United Kingdom (UK), to be able to deliver these treatments in a safe and timely fashion (8). Anxieties around delivery are tied to the potential volume of the task, given that an ageing population means 96 million people are predicted to be living with dementia worldwide by 2050 (9). The FDA label for lecanemab states that the medication is to treat patients with “mild cognitive impairment or mild dementia stage of AD, the population in which treatment was studied in clinical trials” (5). This implies amyloid positivity would need to be determined either by positron emission tomography (PET) imaging or cerebrospinal fluid (CSF) analysis prior to treatment. The label also stipulates that a minimum of four magnetic resonance imaging (MRI) brain scans are required to monitor for side-effects; one in the year prior to initiating treatment and three further scans prior to the 5th, 7th and 14th infusions. Further scans would be indicated if amyloid related imaging abnormalities (ARIA) develop, or symptoms suggestive of ARIA. All of these are resource intensive and would require significant investment in new facilities and staff, on top of the cost of the drug. Currently the National Health Service (NHS) in the UK struggles to meet diagnostic demand, with patients facing a two-year wait in some services (10). Only 36 percent of UK psychiatrists believe their service could adapt to start delivering disease modifying treatments within a year (8).

Attempts have already begun in the USA to manage access to treatment by restricting lecanemab to patients most likely to benefit, in order to manage the financial and infrastructural impact and avoid treatment of patients unlikely to benefit. The USA Department of Veterans Affairs recently approved funding for lecanemab but with “criteria of use” (11). These imposed several inclusion and exclusion requirements for funding, for example excluding individuals under the age of 65 years and those who are homozygous for apolipoprotein E4 (ApoE4). Others have proposed ‘appropriate use recommendations’ (12,13).

Estimates have appeared in the general press of more than 100,000 people potentially being eligible for treatment in the UK, although it is not clear how these figures were derived (14). Others have examined small, specific populations to look at the numbers potentially eligible, but it is not clear how this would translate to a population level (15). In order to plan for delivery of these medicines, the best possible informed estimates of potential demand based on large real-world data sets are urgently needed.

Given this context we set out to use data from large audits and electronic patient record data sources to predict the potential scale of real-world demand as accurately as possible. We considered established criteria for treatment from clinical trials for which we have informative service-level data. We also consider criteria where we have limited, less precise, or no data to guide us but where we can make a reasonable estimate. Though imperfect, we present what we believe to be the best possible estimate at the current time of the maximal numbers of patients who might require treatment in the United Kingdom.

## Method

We used the anonymised research patient record database from Cambridgeshire and Peterborough NHS Foundation Trust (CPFT) for the calendar year 2019. This database has overarching ethical approval from the Cambridge Central Research Ethics Committee (12/EE/0407,17/EE/0442) and this project was further assessed and approved by the CPFT research database committee. The clinical service covers a population of approximately 1 million people, including the population of Cambridgeshire and Peterborough and some surrounding counties. The mental health trust is not the sole provider of memory assessment services. The population is also served by clinics where diagnoses of dementia are made in general hospital, geriatric medicine, and neurology departments, though the latter tend to focus on young-onset disease, or non-Alzheimer dementias such as Huntington’s disease and frontotemporal dementia which would not be within the scope of the treatments discussed here.

In 2019, CPFT was also providing post-diagnostic and emergency support to patients diagnosed in other services resulting in the majority of new diagnoses in the catchment area having had contact with the trust. The imaging data was obtained from our ongoing study Quantitative MRI in NHS Memory Clinics (QMIN-MC, www.rittman.uk/qminmc). This project uses MRI scans obtained during routine NHS memory clinic appointments to develop and evaluate MRI biomarkers for clinical use, including for selection of novel treatments. The scans are all reported by a consultant neuroradiologist. To assess the burden of vascular disease, we calculated the Fazekas score (16) for all QMIN-MC MRI scans collected at our trust.

We evaluated the replicability of our findings using the deidentified patient record from the South London and Maudsley NHS trust (SLaM) (17). SLaM provides comprehensive mental health services to a geographic catchment of around 1.2 million residents in four south London boroughs (Croydon, Lambeth, Lewisham and Southwark) as well as some regional and national specialist services. Although it has a similar population size to Cambridgeshire, it is an urban setting with a much greater ethnic diversity (48% non-White) and most with social deprivation. Cerebrovascular burden was assessed in using the SLaM Image Bank dataset (www.brainregion.com/slamimagebank) and findings were extrapolated to the full SLaM cohort. This was done using free text analysis of the imaging reports to exclude reports with a mention of “moderate small vessel”, “extensive small vessel”, “severe small vessel”, “old infarct”, “superficial siderosis”, “encephalomalacia”, alongside manual reads of reports containing “infarct”, “multiinfarct”, “old infarct”, “haemorrhage”, “microhaemorrhage”, “microhaemorrhages”, “bleed”, “microbleed”, “macrobleed”, “stroke”, “large vessel”, “territory”, “vasogenic oedema”, “oedema”, “lacunar”, “major vascular”, “white matter disease”, “tumour”, “space occupying”, “contusion”, “vascular malformation”, and “aneurysm”. The source data are approved for secondary analysis (Oxford REC, reference 18/SC/0372) (17), and via the National Institute for Health and Care Research (NIHR) BRC Maudsley Neuroimaging Call (NQOD-04) for SLaM Image Bank access (18).

## Results

### Factors influencing numbers requiring treatment where direct data is available in our primary dataset (CPFT)

#### Numbers with an appropriate diagnosis

To avoid data derived from service disruption arising from the Covid-19 pandemic we examined the calendar year 2019. During that year CPFT received referrals for 25,567 unique individuals to mental health services. Of these 7,547 were aged between 50 and 90. Of those, 880 received a new diagnosis of either AD with early or late onset, Alzheimer’s disease atypical or mixed type or Alzheimer’s disease unspecified (ICD-10 F00.0 (16), F00.1 (326), F00.2 (521) or F00.9 (17)). Similarly, 597 received a new diagnosis of mild cognitive impairment (MCI), (ICD-10 F06.7). Forty-five received a diagnosis of MCI and a subsequent diagnosis of dementia in the same year (of which 11 received a diagnosis of vascular dementia), leaving 1432 unique individuals. This is likely to be an overestimate of patients with an appropriate diagnosis as it is questionable whether a patient with a diagnosis of atypical or mixed AD would meet the criteria for entry to the trials of anti-amyloid therapy, and not all those diagnosed with MCI would meet the criteria for MCI secondary to AD. At the very least some 15% of patients with MCI are amyloid biomarker negative (19), meaning the total pool of potential participants would be 1349, which is a parsimonious estimate (i.e., errs on the side of inclusion). Our figure of 15% screen failure for amyloid is less than that seen in some published trials, for example 24% in TRAILBLAZER-ALZ2 (2), and we have only applied it to the MCI group, but throughout where there is a choice of data we have always selected those which predict a higher treatment population so as to identify what might be the highest number of people we might need to accommodate for treatment rather than an underestimate. We have excluded patients on the basis of amyloid positivity at this stage of the calculation, but at what point in the process this is assessed is important in determining the number of PET scans required.

#### Numbers meeting cognitive inclusion criteria

All trials included cognitive criteria for entry. CLARITY-AD used the mini-mental state examination (MMSE) combined with age-adjusted Wechsler memory scale IV – Logical Memory II, a measure of cognition not commonly employed in NHS clinical services (1). TRAILBLAZER-ALZ used the MMSE alone, excluding patients whose MMSE was not in the range 20-28. MMSE scores can be converted to an estimated Addenbrooke’s Cognitive Examination-Revised (ACE-R (20)) score, which is similar to the Addenbrooke’s Cognitive Examination-III (ACE-III (21)) test used in our clinics. An MMSE score of 20 equates to an ACE-R score of 52 (22). 90% of patients in our service who had an ACE administered scored more than 52. This is again likely to be a conservative estimate as patients presenting late in the disease course who are very impaired would not be tested with an ACE but instead with a more basic tool, such as a the mini-ACE. An ACE score of 52 is low and likely to include patients who would not meet the criteria for ‘early’ disease. Nevertheless, using the most parsimonious assumption that 90% have an appropriate cognitive score leaves a potential treatment population of 1214.

#### Numbers able to undergo imaging

Eligibility criteria for all trials of anti-amyloid therapy include confirmation of amyloid and sometimes tau positivity using PET or CSF, as well as structural imaging to exclude a significant burden of vascular disease. Our patients do not routinely undergo CSF examination or amyloid and/or tau PET imaging, but MRI scanning is part of the routine diagnostic work-up. We have previously published data showing that in our local service only 66% of patients are imaged (including those patients who had recently been imaged for other reasons) (23). The 2019 national memory service audit found the average proportion of patients in each service deemed not to need imaging was 15% (24). If 85% of patients undergo appropriate structural imaging, this leaves 1032 of our population remaining potentially eligible for treatment.

#### Numbers who meet criteria for vascular burden

Our routinely collected diagnostic scans in CPFT are anonymised and available for research purposes. We examined this library to see what percentage of patients might be excluded using the imaging criteria for CLARITY-AD i.e., ‘Other significant pathological findings on the MRI scan including but not limited to: more than 4 microhaemorrhages…; a single macro haemorrhage…; an area of superficial siderosis, evidence of vasogenic edema, evidence of cerebral contusion, encephalomalacia, aneurysms, vascular malformations or infective lesions, evidence of multiple lacunar infarcts or stroke involving a major vascular territory, severe small vessel or white matter disease, space occupying lesions or brain tumors…’ (1). Of 100 available scans stored for research purposes with a diagnosis of MCI or Alzheimer’s disease, 49% had a total Fazekas score of four or more and 40% had a deep white matter lesion score of 2 or more. Applying the latter of these two figures decreases the total potential patient pool for treatment to 619.

#### Numbers excluded for other reasons

Other exclusion criteria from CLARITY-AD included pregnancy, neurological conditions adding to cognitive impairment beyond AD, history of TIA stroke or seizures within 12 months of screening, psychiatric diagnosis that could interfere with treatment, geriatric depression scale >-8, concomitant treatment with biological treatment, bleeding disorders, B12 or TSH abnormalities, HIV positivity, malignant neoplasms within 3 years of screening, suicidal ideation, other abnormalities, drug or alcohol abuse, planned surgery, severe sensory impairment or participation in other trials was either not known and/or not thought likely to be as a significant contributor to limiting treatment (1).

We examined the data for co-morbid ICD10 diagnostic F codes (mental, behavioural and neurodevelopmental disorders). 68 patients had a recorded diagnosis of delirium, 17 depressive disorder and 13 mixed anxiety and depression. All other psychiatric diagnostic categories yielded less than 10 individuals. Though the additional exclusion criteria might reduce the potential treatment population further, we do not have data to address all of them and it seems unlikely any one would exclude a large number of patients, so we did not adjust the total potential treatment population.

### Factors influencing numbers requiring treatment where indirect data is available

Further questions arise as to how many of those eligible would be willing to undergo the requirements of treatment. These include repeat imaging and a fortnightly or monthly intravenous infusion with significant risk of side effects. A parsimonious approach might be to look at how many patients with a diagnosis of AD currently receive oral symptomatic treatments such as donepezil and memantine. Previously we showed that 65% of eligible patients are prescribed these medicines in our service, comparable to larger data sets such as the national 2019 memory service audit, which found that 83% of eligible patients were offered currently available medication, of which 90% accepted, meaning 75% of the overall sample received treatment (23,24). Applying the more generous 75% cut off leaves a pool of 464 patients to receive treatment. Of course, it may be patients would be more willing to take a disease modifying treatment which would increase numbers or that contraindications to existing treatments may not apply to new ones, but it is probably more likely that the significant extra burden of imaging, intravenous treatment and safety scans means 75% is a number which errs strongly on the side of inclusion.

Using real data from the service in Cambridgeshire but taking a parsimonious view of patient selection at every point to give the greatest possible treatment population results in a total maximum potential treatment population of 464 from a total number of new diagnoses of 1432. This is in the context of the 70% screen failure rate in CLARITY-AD (which if applied to all new diagnoses in our service would suggest a potential treatment population of 430) and 87% in TRAILBLAZER-ALZ (potential treatment population of 186). Based on our data we might expect a maximum of 464 people starting treatment in Cambridgeshire per year.

The duration of therapy beyond the 18 months for which there is an established evidence base remains unclear. To date, no stopping criteria for anti-amyloid therapy have been described, although there is a suggestion that treatment should not be continued once patients develop moderate dementia (defined as progression to CDR global score of 2.0, MMSE score below 20, and loss of autonomy in key ADLs) due to lack of evidence of efficacy at this stage of the condition (13). Patients in the TRAILBLAZER-ALZ 2 trial stopped infusion when amyloid was supressed below a threshold, meaning around a third ceased treatment after six months (2). Patients with Alzheimer’s disease spend between 2 and 6 years in the mild dementia stage, which depends on factors such as sex, age at diagnosis and ApoE4 status (25). Amyloid burden and the occurrence of ARIA may also influence when treatment is stopped. Ongoing clinical trials will hopefully shed light on whether maintenance therapy is required, or whether treatment can stop when patients become amyloid negative. The majority of ARIA-E (oedema or effusions) occurred in the first few months of lecanemab treatment (71% occurred within three months (1)), so this is unlikely to contribute to later dropouts. However, isolated ARIA-H (cerebral microhaemorrhages, macrohaemorrhages, or superficial siderosis) occurred more randomly during the treatment course, and at a similar rate for the lecanemab (8.9%) and placebo (7.8%) groups (1). If future stopping criteria includes the development or extent of ARIA-H, then this will increase the number of people who drop out. In CLARITY-AD, patients were treated with lecanemab for 18 months and had a 10% per year all cause drop out; this rate is likely to be higher in a clinical setting than in a trial (1).

If we assume 464 patients starting treatment each year, and a maximal mean treatment period of three years with infusions occurring fortnightly, then the numbers requiring treatment would reach steady state at year three with 1,258 individuals needing 629 treatments per week or 90 per day for Cambridgeshire and Peterborough. The numbers of infusions would be halved in the event of monthly treatment and lower still if treatment is stopped once amyloid is suppressed.

### Factors influencing numbers requiring treatment where no data is available

There are a number of further factors which are impossible to know or use data to predict. One is whether the advent of potentially disease modifying treatment might encourage more people to present for assessment. If that were to happen then memory clinics in the UK struggling to meet current demand would need scaling up to meet this increased need within a reasonable timeframe. For example, in the most recent UK national memory assessment audit the average time to diagnosis was over 17 weeks, with a range of 0 to 104 weeks (10). If patients move from a point in disease beyond that where treatment is possible whilst waiting this will not be acceptable. Availability of these treatments may increase willingness to undergo investigation, such as brain scans, with attendant pressure on these services. Further unknowns are whether recent advances in blood biomarkers might obviate the need for PET imaging or CSF analysis for entry, or whether the advent of a treatment administered subcutaneously might increase demand. It is possible decisions around licensing and access may be influenced by data from trials of subgroups of patients. For example, consideration has been given to whether funding might be restricted for ApoE4 homozygotes, where the balance of risk and benefit might differ, which would screen out a further 2% of the population (11). Lastly, we have considered the use of these medicines only within the criteria used in their clinical trials as it is important to use them only in the context within which they have been shown to be of benefit. However, there is a history of drugs being used more widely post-licensing with consequent increased use and resource requirement.

### Replication

In order to validate our findings in a second independent dataset, we applied the same assumptions to the South London and Maudsley (SLaM) NHS Trust anonymised patient record and SlaM Image Bank dataset (18). In the year 2019 SlaM received 56,819 referrals, of which 11,489 were aged 50-90. Of these 1,880 received a diagnosis which would qualify them for treatment. After removing 15% of patients with MCI assumed to be amyloid-negative, 876 met the threshold in cognitive tests and 745 were expected to undergo neuroimaging. Within the cohort, 115 had neuroimaging available for analysis, with 24 patients (21%) being excluded on the basis of significant vascular burden. Extrapolating this to the full SlaM cohort leaves a total potential treatment population of 589. If 75% accept treatment, the number starting treatment each year would be 442, a similar number to those starting treatment in Cambridgeshire. The results from the two trusts are summarised in Figure 1. Taken together, these two trusts cover around 2.2 million (3%) of the UK’s total population of 67 million. Therefore, our calculations suggest an annual demand of 30,200 people starting treatment each year in the UK, a much lower number than previous estimates reported in the press. Whilst this is the potential treatment population, in reality initial treatment uptake is likely to be slow and patchy as patients, clinicians and services adapt to any new treatments becoming available with use slowly increasing over time.

**Figure 1.**
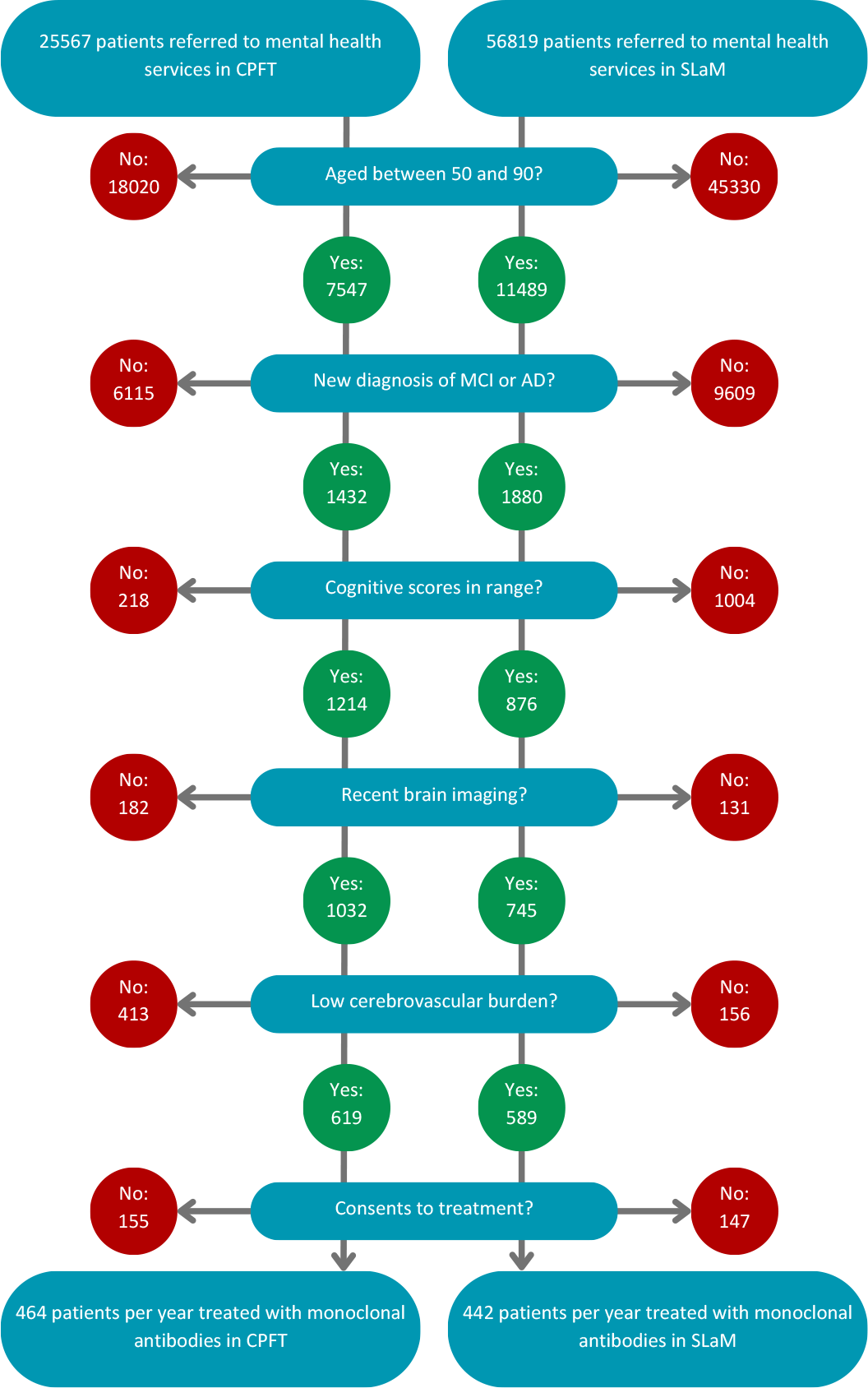
Flow diagram showing the number of patients in Cambridgeshire and Peterborough NHS Foundation Trust (CPFT) and South London and Maudsley NHS Trust (SLaM) who would receive treatment with monoclonal antibodies for Alzheimer’s disease.

## Discussion

We have used available data to make the best possible prediction of maximal potential demand for new AD treatments for two large NHS trusts in the UK, which we have extrapolated nationally. These figures are based on hard data as far as possible, though inevitably they also require some assumptions. Where assumptions have been made, we have attempted to explain our careful reasoning and the calculation can be easily adjusted using different parameters as new information comes to light or if people choose to make different assumptions. The numbers may cause concern for services already struggling to meet current demand. However, it is worth making a comparison with use of monoclonal antibodies for other conditions. According to the local prescribing data for Cambridgeshire and Peterborough, 432 new patients were treated with monoclonal antibodies for inflammatory bowel disease in 2022. Although this is similar to our calculated demand for Alzheimer’s disease, most monoclonal antibodies are administered subcutaneously less frequently than every two weeks, for shorter periods of time, and do not need MRI monitoring for side-effects. Nevertheless, the challenge posed by monoclonal antibodies for Alzheimer’s disease is not out of keeping in terms of scale with these agents being used in other conditions.

One significant issue might be the availability of structural neuroimaging prior to treatment. National Institute for Health and Clinical Excellence (NICE) guidance (26) for the diagnosis of dementia recommends neuroimaging but does not specify the modality, i.e. MRI or computed tomography (CT). National audits suggest 74% of NHS memory services use CT rather than MRI scans, so routine availability of MRI scans will limit access to treatment (24). This would be further underlined if treatment requires serial MRI scanning after initiation, which seems likely with a minimum of four scans recommended in the US. The potential need for PET scanning to identify appropriate patients for treatment would likely be an even greater issue. The number of PET scans required would depend where on the pathway to determine eligibility they are placed, but even if applied only to those that have met all other inclusion criteria this would be hundreds of PET scans per year regionally and tens of thousands nationally as the number of scans for eligibility will be greater than the number of those eligible for treatment by up to 25 % (2). These requirements for imaging would be challenging, perhaps even impossible, given current resources. This difficulty might be ameliorated by the use of CSF examination (which would itself provide new issues) or blood biomarkers to identify patients, but it would currently present a rate limiting step in access. The phase three trial results also suggest an altered risk of ARIA depending on ApoE genotype (1,2). It seems likely that this genetic information would be helpful in informing patients and scaling up the facility for testing and appropriate counselling would present a further challenge to health services not currently routinely providing this. ApoE genotyping is already included in some ‘appropriate use’ recommendations (12,13).

Whether these drugs or other monoclonal antibodies for AD are licensed in the UK, approved by NICE and accepted by patients based on the balance of benefits and risks remains to be seen. If approved they would undoubtedly present a significant challenge to deliver, particularly given existing service pressures, but the challenge would not be completely out of keeping with the delivery of similar biologic treatments for other conditions such as inflammatory bowel disease, cancer or multiple sclerosis where new treatments have been shown to be cost-effective. Although the clinical benefit of the current generation of anti-amyloid drugs appears marginal, if they are licensed and can demonstrate cost-effectiveness, which must include the costs to healthcare systems of setting up and maintaining the delivery infrastructure as well as the price of the drug itself, then they should be made available to patients on an equitable basis. We believe the data presented here is the best estimate of potential demand to date, and we hope that it will support healthcare planners and policymakers in preparing for any future delivery of these drugs.

## Data Availability

The data that support the findings of this study are available from the corresponding author upon reasonable request.

## Declaration of interests

B.R.U. has been principal investigator in a number of commercial trials of novel agents in Alzheimer’s disease, including anti amyloid therapy, though has not been involved in any of the trials referenced here. He is a member of the Faculty of Old Age Psychiatry Executive Committee. He has received ad hoc payments for advisory roles to market research companies representing pharma. He is the R&D director at his trust (which has received financial support from industry to develop its clinical trials infrastructure), CRN lead for dementia for the east of England and national CRN lead for stratified medicine in dementia.

R.B. has worked on Roche studies (including CI for Graduate study with gantenerumab) and Biogen (sub-investigator for aducanumab) and has previously received ad hoc payments from Roche and Biogen for advisory roles.

J.D.I. is clinical director of NHS England (London) dementia clinical network. He has received conference expenses and consultancy fees (paid to his institution) from Roche, a speaker’s fee (paid to his institution) from Biogen and payment (to his institution) from Nestle Health Science for membership of a clinical trial steering committee.

A.V.V. has received grants from the Alzheimer’s Society, Alzheimer’s Research UK, and NIHR BRC, including an NIHR BRC Maudsley Neuroimaging Grant. A.V.V. is funded by NIHR as NIHR Clinical Lecturer and supported by the NIHR Maudsley Biomedical Research Centre at South London and Maudsley NHS Foundation Trust and King’s College London (NQOD-04) as PI of SLaM Image Bank.

C.M. is part-funded by NIHR Biomedical Research Centre at South London and Maudsley NHS Foundation Trust and King’s College London.

R.S. declares research support received in the last 3 years from Janssen, GSK and Takeda.

M.S.K. has attended as a panel member at ARUK conference on new treatments.

A.A.S.L. is funded by the NIHR as an Academic Clinical Fellow and is a member of the NIHR Dementia Portfolio Development Group.

T.R., T.S., C.M., M.M., J.L. have nothing to declare.

## Funding Statement

This research, and the QMIN-MC study, was supported by the NIHR Cambridge Biomedical Research Centre (NIHR203312). The views expressed are those of the author(s) and not necessarily those of the NIHR or the Department of Health and Social Care. B.R.U.’s post is part funded by a generous donation from Goldman Sachs giving. R.S. is part-funded by: i) the NIHR Biomedical Research Centre at the South London and Maudsley NHS Foundation Trust and King’s College London; ii) the NIHR Applied Research Collaboration (ARC) South London at King’s College Hospital NHS Foundation Trust; iii) UKRI – Medical Research Council through the DATAMIND HDR UK Mental Health Data Hub (MRC reference: MR/W014386); iv) the UK Prevention Research Partnership (Violence, Health and Society; MR-VO49879/1), an initiative funded by UK Research and Innovation Councils, the Department of Health and Social Care (England) and the UK devolved administrations, and leading health research charities.

For the purpose of open access, the author has applied a Creative Commons Attribution (CC BY) licence to any Author Accepted Manuscript version arising from this submission.

The replication of this project is approved under CRIS Ethics (Oxford REC, reference 18/SC/0372), sub-project ID CRIS 21-064, and part of the NIHR BRC Maudsley Neuroimaging Call (NQOD-04) for SLaM Image Bank.

## Author contributions

Design of the study (B.R.U.). Data collection and analysis (B.R.U., T.R., T.S., M.M., J.L., C.M., A.A.S.L., A.V.V.). Drafting the manuscript (B.R.U., A.A.S.L., A.V.V., T.R., J.D.I., M.S.K., R.B., C.M., R.S.). Revision of the manuscript (B.R.U., A.A.S.L., A.V.V.).

## Notes

### Author Declarations

We used the anonymised research patient record database from Cambridgeshire and Peterborough NHS Foundation Trust (CPFT) for the calendar year 2019. This database has overarching ethical approval from the Cambridge Central Research Ethics Committee (12/EE/0407,17/EE/0442) and this project was further assessed and approved by the CPFT research database committee. We evaluated the replicability of our findings using the deidentified patient record from the South London and Maudsley NHS trust (SLaM) The source data are approved for secondary analysis (Oxford REC, reference 18/SC/0372), and via the National Institute for Health and Care Research (NIHR) BRC Maudsley Neuroimaging Call (NQOD-04) for SLaM Image Bank access.

